# Derivation of an electronic frailty index for short-term mortality in heart failure: a machine learning approach

**DOI:** 10.1101/2020.12.26.20248867

**Authors:** Chengsheng Ju, Jiandong Zhou, Sharen Lee, Martin Sebastian Tan, Ying Liu, Yuhui Zhang, Tong Liu, Esther WY Chan, Ian Chi Kei Wong, Li Wei, Qingpeng Zhang, Gary Tse

**Affiliations:** Research Department of Practice and Policy, School of Pharmacy, University College London, London, United Kingdom; School of Data Science, City University of Hong Kong, Hong Kong, Hong Kong SAR, China; Laboratory of Cardiovascular Physiology, LKS Institute of Health Sciences, Chinese University of Hong Kong, Hong Kong SAR, China; Faculty of Arts and Science, University of Toronto, Toronto, Canada; Heart Failure and Structural Cardiology Division, Department of Cardiology, The First Affiliated Hospital of Dalian Medical University, Dalian, Liaoning Province, 116011, China; Heart Failure Center, State Key Laboratory of Cardiovascular Disease, Fuwai Hospital, National Center for Cardiovascular Diseases, Chinese Academy of Medical Sciences & Peking Union Medical College, Beijing, China; Tianjin Key Laboratory of Ionic-Molecular Function of Cardiovascular disease, Department of Cardiology, Tianjin Institute of Cardiology, Second Hospital of Tianjin Medical University, Tianjin 300211, China; Centre for Safe Medication Practice and Research, Department of Pharmacology and Pharmacy, The University of Hong Kong, Hong Kong SAR, China; Faculty of Health and Medical Sciences, University of Surrey, GU2 7AL, Guildford, United Kingdom

**Keywords:** Frailty index, heart failure, mortality, inflammation, nutrition, machine learning

## Abstract

**Objective:** Frailty may be found in heart failure patients especially in the elderly and is associated with a poor prognosis. However, assessment of frailty status is time-consuming and the electronic frailty indices developed using health records have served as useful surrogates. We hypothesized that an electronic frailty index developed using machine learning can improve short-term mortality prediction in patients with heart failure.

**Methods:** This was a retrospective observational study included patients admitted to nine public hospitals for heart failure from Hong Kong between 2013 and 2017. Age, sex, variables in the modified frailty index, Deyo’s Charlson comorbidity index (≥2), neutrophil-to-lymphocyte ratio (NLR) and prognostic nutritional index (PNI) were analyzed. Gradient boosting, which is a supervised sequential ensemble learning algorithm with weak prediction submodels (typically decision trees), was applied to predict mortality. Comparisons were made with decision tree and multivariate logistic regression.

**Results:** A total of 8893 patients (median: age 81, Q1-Q3: 71-87 years old) were included, in whom 9% had 30-day mortality and 17% had 90-day mortality. PNI, age and NLR were the most important variables predicting 30-day mortality (importance score: 37.4, 32.1, 20.5, respectively) and 90-day mortality (importance score: 35.3, 36.3, 14.6, respectively). Gradient boosting significantly outperformed decision tree and multivariate logistic regression (area under the curve: 0.90, 0.86 and 0.86 for 30-day mortality; 0.92, 0.89 and 0.86 for 90-day mortality).

**Conclusions:** The electronic frailty index based on comorbidities, inflammation and nutrition information can readily predict mortality outcomes. Their predictive performances were significantly improved by gradient boosting techniques.

## Introduction

Frailty refers to a reduced physiological reserve leading to an impairment in resilience from physical distress. Compared to highly functional community-dwelling elders, frail older adults are more likely to experience falls and disability, contributing to frequent hospitalization and premature death (1). Conventional evaluation of frailty relies on physical examination. However, this precludes its calculation using administrative data such as electronic health records. Recently, a claims-based frailty scoring system has been validated against Fried and colleges’ frailty phenotype using a claim database in the United States (2-4). These electronic frailty indices do not normally include measures of chronic inflammation or nutrition status, which are both closely related to frailty syndrome and are strong determinants of adverse outcomes such as mortality (5, 6).

Heart failure is a complex syndrome characterized by high prevalence in older patients and poor prognosis (7). Heart failure and frailty have an overlapping phenotype and their co-existence is common (8). Given these associations, there has been several studies exploring the intersections between heart failure and frailty (8). Importantly, frailty has been recognized as a major prognostic indicator of heart failure, in which patients with concurrent frailty and heart failure have increased risks of hospitalizations and mortality (9).

Furthermore, inflammation and nutrition status are known independent predictors of heart failure outcomes (10, 11). Inflammation has a pivotal role in the pathogenesis of heart failure. It can trigger cardiac remodeling and dysfunction that further induce cardiomyocyte damage that underlies heart failure (12). Moreover, it has been proposed that co-morbidities, such as diabetes and obesity, can induce a systemic pro-inflammatory state that drives the myocardial structural and functional alterations in heart failure (13). Conversely, inflammation can also be a consequence of established heart failure via the mechanisms of increased wall stress on endothelial cells, cell death and oxidative stress (ROS) (14). In this regard, inflammation and heart failure are interconnected and mutually inducing. Elevated pro-inflammatory cytokines were found to associate with worse clinical outcomes in patients with heart failure (15, 16), and some studies have demonstrated that NLR can be used as a prognostic factor for heart failure (17). Similar to inflammation, malnutrition is also an independent risk factor and prognostic factor for heart failure (18, 19). Various nutritional metrics, including PNI, associate well with the survival outcomes (20, 21). Both factors possess important predictive values for clinical outcomes in patients with heart failure (22-24). Despite the multiple associations between frailty, heart failure, nutrition status and inflammation, whether incorporating the measures of nutrition status and inflammation into the existing frailty index can enhance its predictive value on outcomes of heart failure remains unknown.

Machine learning techniques has gained popularity in medical research. Specifically, gradient boosting has recently been explored as a method to predict adverse outcomes in heart failure (25). In this study, using a large cohort of patients with heart failure, we tested the hypothesis that incorporating neutrophil-to-lymphocyte ratio (NLR) and prognostic nutritional index (PNI) into an electronic frailty index using gradient boosting, a machine learning approach will improve prediction for short-term mortality risks.

## Methods

### Data source and study population

The territory-wide retrospective study was approved by The Joint Chinese University of Hong Kong - New Territories East Cluster Clinical Research Ethics Committee. This is a retrospective cohort study nested within the territory-wide Clinical Data Analysis and Reporting System (CDARS), an electronic health record system managed by the Hong Kong Hospital Authority since 1995. The database included over 7 million Hong Kong residents and has been used for producing high-quality clinical studies (26, 27). Patient information was de-identified with pseudo-identity numbers. Clinical data available from CDARS includes demographic information, diagnosis, procedure, prescription, laboratory test results, admission/discharge information and death information.

The inclusion criterion was patient admitted to any of the nine local hospitals during a four-year period between July 2013 and July 2017 with a principal diagnosis of heart failure. The diagnosis of heart failure was defined as having a record with the International Classification of Diseases, Ninth Revision, Clinical Modification (ICD-9 CM) codes of 428.X.

### Study variables

Variables that were previously included in the modified frailty index (4) were identified from the relevant ICD-9 codes. These include depression, Parkinson’s disease, arthritis, paranoia, chronic skin ulcer, pneumonia, falls, skin and subcutaneous tissue infection, mycoses, gouty arthropathy and urinary tract infection. Laboratory test results on the measures of albumin level, neutrophil and lymphocyte counts were extracted to calculate inflammatory and nutritional indices. Neutrophil-to-lymphocyte ratio (NLR) was given by the ratio of peripheral neutrophil count/mm^3^ to peripheral lymphocyte count/mm^3^. Prognostic nutritional index (PNI) was calculated by 10 × serum albumin value (g/dl)□+□0.005 × peripheral lymphocyte count/mm^3^. NLR and PNI estimates nearest to the admission time of the first heart failure related hospitalization of the patients were used in the analysis. Baseline Deyo’s Charlson comorbidity index incorporating 17 major medical conditions was also included as a single score (28). A comparison of the included variables used in the modified frailty index, Charlson Deyo’s Charlson comorbidity index and our electronic frailty index is shown in **Supplementary Appendix Table S1**.

### Outcomes and statistical analysis

The primary outcomes were 30-day and 90-day mortality, from the date of the first heart failure related-hospitalization of the patients. The outcome of 30-day mortality is binary and equals to 1 for mortality within 30 days and 0 otherwise, and the same for 90-day mortality outcome. Continuous variables were presented as median (interquartile range [IQR]) and categorical variables were presented as count (%). The Mann-Whitney U test was used to compare continuous variables. The χ^2^ test with Yates’ correction was used for 2×2 contingency data, and Pearson’s χ^2^ test was used for contingency data for variables with more than two categories. To identify significant risk factors associated with 30-day and 90-day mortality, univariate logistic regression was used to determine odds ratios (ORs) and 95% CIs. Significant variables from the univariate logistic regression (p<0.05) were further included in multivariate logistic regression to build the frailty model.

The idea of frailty is the cumulative deficits, each of which in isolation may not exert significant effects. To test this idea, we conducted an additional multivariate logistic regression analysis incorporating all risk variables, including the non-significant variables from univariate logistic regression. Finally, to demonstrate the utility of NLR and PNI, both variables were excluded in sensitivity analysis to examine the effects on evaluation metrics.

A two-sided α of less than 0.05 was considered statistically significant. All statistical analyses were performed using RStudio software (Version: 1.1.456).

### Machine learning model development

Gradient boosting is a typical type of machine learning boosting, relying on the intuition that the best possible next model, when sequentially combined with previous weak models (e.g. decision trees) in a stage-wise fashion, is able to minimize the overall prediction error measured by performance evaluators, e.g., precision, recall, the area under the curve (AUC). Weaker learning models are fitted through loss gradient minimization with gradient descent optimization algorithm (29). This method was used for mortality prediction in heart failure based on administrative claims with electronic health records (30). Variable importance ranking was generated to construct a machine learning based risk score for mortality prediction. Partial dependence plots were provided as low-dimensional graphical renderings of marginal effects to assist in the interpretation of relationships between most important variables and the mortality outcome. A Five-fold cross validation was performed to compare the performance in terms of precision, recall and area under the curve (AUC) of the gradient boosting model with decision tree model and logistic regression model. The R packages, *gbm* (Version 2.1.5) and *ggplot2* (Version 3.3.2), were used to generate the mortality prediction results.

## Results

In our HF cohort (n = 8893), the median age was 81 (IQR 71-87) years and 45% (n=4027) were males. The baseline characteristics, individual variables included in the modified frailty index, inflammatory and nutritional indices between the patients died within 90 days and the patients without 90-day mortality are shown in **Table 1**. The median cell counts for lymphocytes was 1.2*10^9^/L and for neutrophils was 5.4*10^9^/L, yielding a neutrophil-to-lymphocyte ratio (NLR) of 4.4 (IQR 2.7-7.8). Albumin took a median level of 37.8g/L, yielding a prognostic nutritional index of (IQR 39.8-48.5) (PNI, given by 10 × serum albumin value (g/dl)□+□0.005 × peripheral lymphocyte count (per mm^3^)).

**Table 1.** Baseline characteristics of the heart failure cohort

|  | 90-day mortality<br>N=1472 | No mortality<br>N=7421 | P-value |
| --- | --- | --- | --- |
| **Gender** |  |  |  |
| Male (%) | 639 (43.4%) | 3388 (45.7%) | 0.114 |
| **Age** | 84.4 [77.5-90.1] | 80.0 [70.1-85.9] | **<0.001** |
| **Modified Frailty Index** |  |  |  |
| Depression | 1 (0.1%) | 15 (0.2%) | 0.276 |
| Parkinson's disease | 18 (1.2%) | 55 (0.7%) | 0.061 |
| Arthritis | 7 (0.5%) | 33 (0.4%) | 0.872 |
| Paranoia | 0 (0.0%) | 7 (0.1%) | 0.239 |
| Skin ulcer | 35 (2.4%) | 71 (1.0%) | **<0.001** |
| Pneumonia | 549 (37.3%) | 1492 (20.1%) | **<0.001** |
| Falls | 36 (2.5%) | 154 (2.08%) | 0.369 |
| Skin and soft tissue infection | 17 (1.2%) | 82 (1.1%) | 0.868 |
| Mycoses | 2 (0.1%) | 17 (0.2%) | 0.479 |
| Gouty arthropathy | 87 (5.9%) | 354 (4.8%) | 0.066 |
| UTI | 209 (14.2%) | 560 (7.6%) | **<0.001** |
| Charlson score≥2 | 775 (52.7%) | 697 (47.4%) | **<0.001** |
| **Inflammatory and nutritional indices** |  |  |  |
| PNI | 41.5 [37.0-46.0] | 44.5 [40.4-48.8] | **<0.001** |
| NLR | 5.5 [3.1-9.8] | 4.2 [2.6-7.4] | **<0.001** |

UTI, urinary tract infection; PNI, prognostic nutritional index; NLR, neutrophil-to-lymphocyte ratio.

### Predictors of adverse outcomes and frailty model

Of the 8893 patients with HF, 758 patients died within 30 days (9%) and 1472 died within 90 days (17%) of admission. The findings of univariate logistic regression are reported in **Table S2**. Age, chronic skin ulcers, pneumonia, urinary tract infection, NLR and PNI were significant predictors of 30-day mortality (**Table S2**, left). For 90-day mortality, the same variables that predicted 30-day mortality, as well as Charlson score ≥2 were significant predictors (**Table S2**, right). Subsequently, the significant variables from the univariate analysis were included in multivariate logistic regression. The results of multivariable logistic regression for 30-day and 90-day mortality prediction with all variables are reported in **Table S3** and **Table S4**, respectively. Age, pneumonia, UTI, PNI, and NLR remained significant predictors of both 30-day and 90-day mortality (P<0.05).

### Gradient boosting learning results and frailty score

Five-fold cross validation experiments were conducted with gradient boosting learning. The key to gradient boosting is to set the target outcomes to minimize the overall error in relation to precision, recall and AUC. In this way, the gradient boosting model sequentially adds weak decision tree learning models to the ensemble where subsequent models correct the prediction errors of prior models (**Figure S1**), from which we can see that probability of 30-day mortality and 90-day mortality increases drastically as age grows above 80 years old. Specifically, predictions given by the sequential models that are close to the actual outcome should reduce the overall error, and the process continues until minimized total prediction error is achieved.

A total of 1400 and 1500 trees for 30-day and 90-day mortality prediction were assigned, respectively. The optimal tree number was identified using sensitivity analysis by plotting the value of the out-of-bag (OOB) error rate according to the number of trees within the forest (**Figure S2**). OOB samples are those samples that are not included in the bootstrap samples. Original training data is randomly sampled-with-replacement generating small subsets of data, also known as bootstrap samples. These bootstrap samples are then fed as training data to the forest model. The OOB approach was used for selecting the optimal tree number of the forest model in which four-fifths (as training) of the data was used for constructing the predictive classifier while the remaining was used for evaluating the performance of the forest model. Tree depth was set at 1 for both 30-day and 90-day mortality prediction according to tree depth parameter tuning (**Figure S3**). The variable importance is reported in **Table 2** and is used for building the frailty score.

**Table 2.** Variable importance for 30-day and 90-day mortality prediction with gradient boosting learning

| **30-day mortality** |  | **90-day mortality** |  |
| --- | --- | --- | --- |
| **Variable** | **Importance** | **Variable** | **Importance** |
| PNI | 37.45 | Age | 36.25 |
| Age | 32.11 | PNI | 35.28 |
| NLR | 20.46 | NLR | 14.59 |
| Pneumonia | 6.04 | Pneumonia | 7.05 |
| Skin ulcer | 1.16 | UTI | 2.29 |
| UTI | 1.03 | Skin ulcer | 1.43 |
| Parkinson's disease | 0.49 | Male sex | 0.57 |
| Male sex | 0.45 | Falls | 0.48 |
| Skin and soft tissue infection | 0.25 | Parkinson's disease | 0.46 |
| Gout | 0.23 | Charlson score≥2 | 0.46 |
| Falls | 0.19 | Arthritis | 0.35 |
| Charlson Score≥2 | 0.13 | Skin and soft tissue infection | 0.34 |
| Arthritis | 0.01 | Gout | 0.31 |
| Mycoses | 0.01 | Mycoses | 0.15 |
| Depression | 0 | Depression | 0 |
| Paranoia | 0 | Paranoia | 0 |

UTI, urinary tract infection; PNI, prognostic nutritional index; NLR, neutrophil-to-lymphocyte ratio.

The partial dependence relationships of the highest variable importance values for mortality prediction were also identified using gradient boosting learning. The probabilities of 30-day and 90-day mortality both increase as patient becomes older, and it increases sharply when patients are older than 80 **(Figure 1)**. For PNI, the likelihood of mortality decreases sharply as PNI increases from 0 to 20 and remains almost constant when PNI increases beyond 65 (30-day mortality) or 70 (90-day mortality) (**Figure 2)**. There is a non-linear relationship between NLR and 30- and 90-day mortality (**Figure 3**). Patients with pneumonia has high probability of mortality, 24% for 30-day mortality and 14% for 90-day mortality.

**Figure 1.**
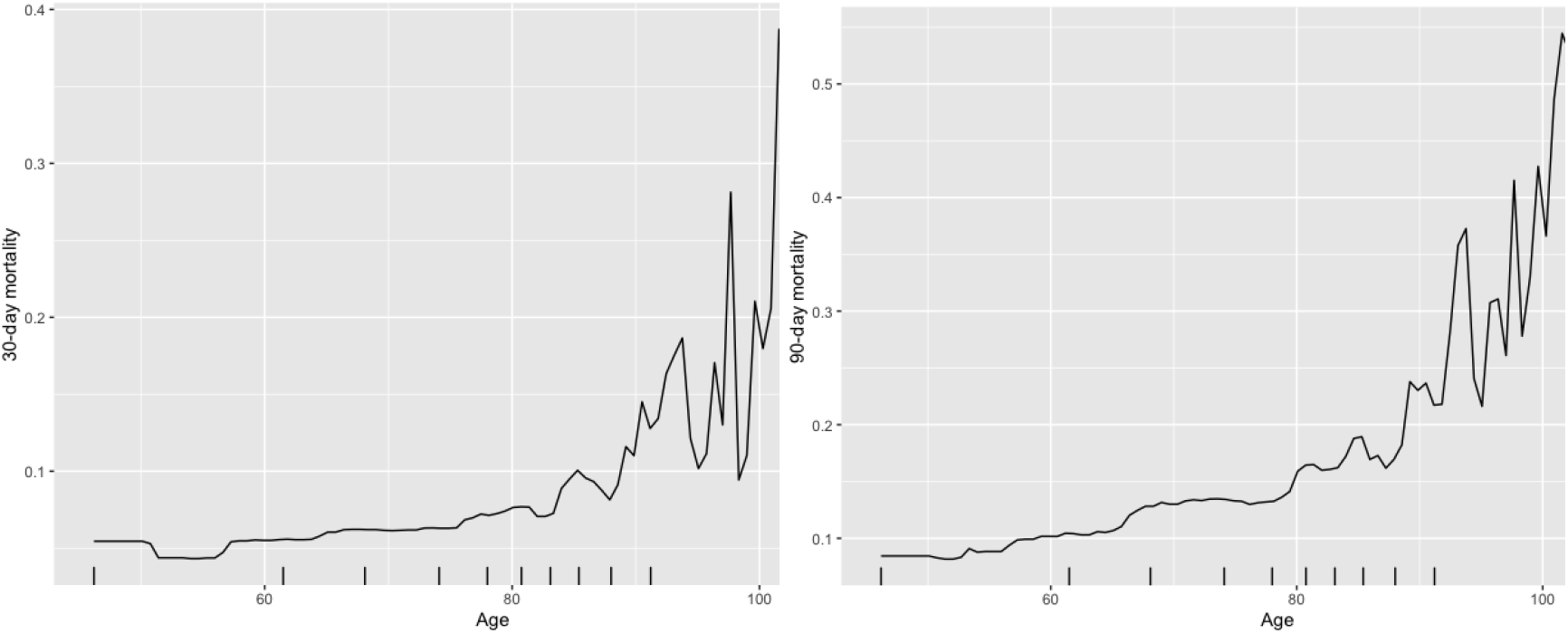
Partial dependence of patient age for 30-day (left) and 90-day (right) mortality risk probability prediction.

**Figure 2.**
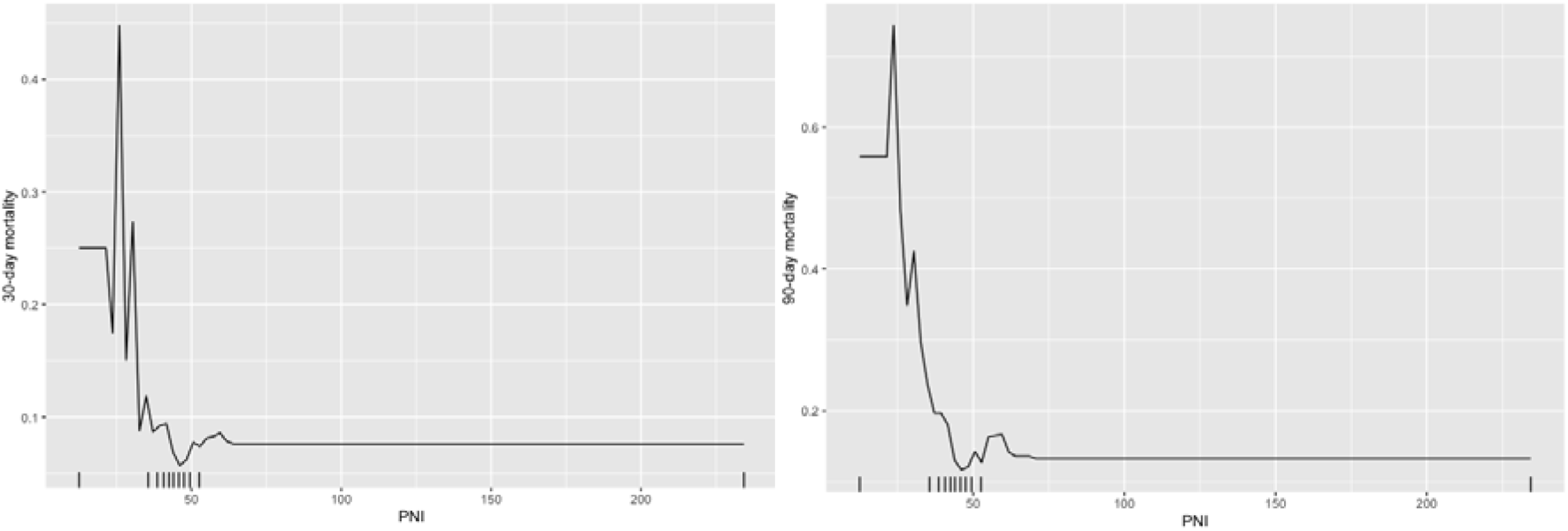
Partial dependence of PNI for 30-day (left) and 90-day (right) mortality risk probability prediction.

**Figure 3.**
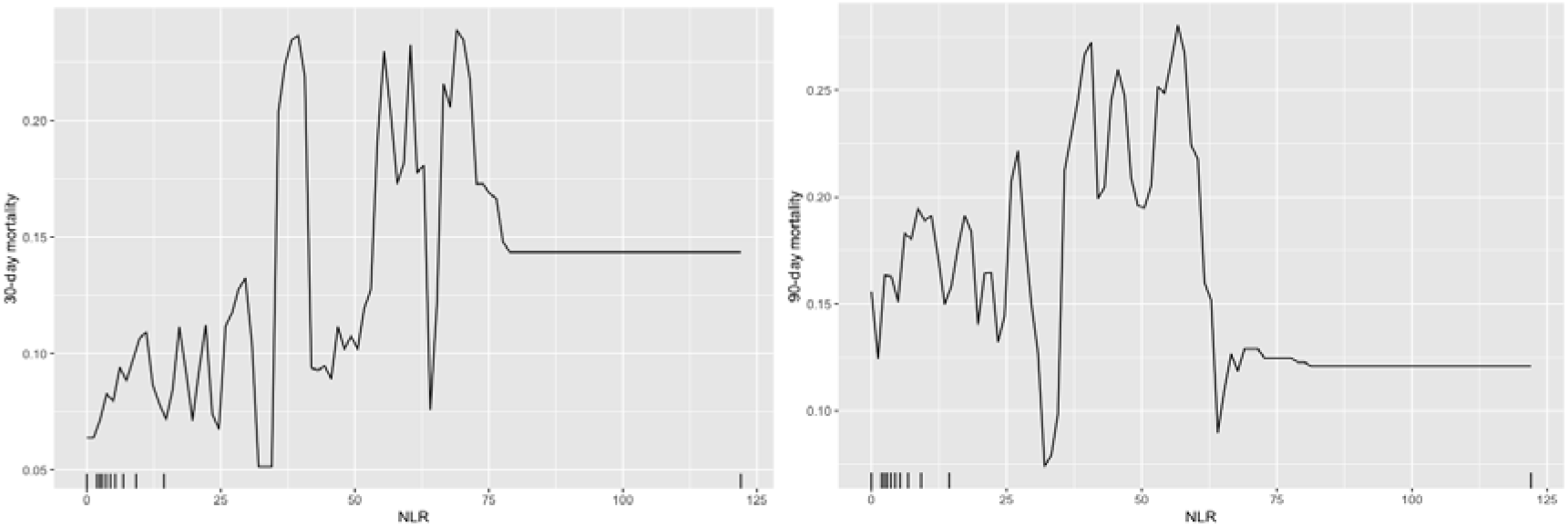
Partial dependence of NLR for 30-day (left) and 90-day (right) mortality risk probability prediction.

Comparative analyses of gradient boosting learning model, decision tree model, and multivariate logistic regression model for 30-day and 90-day mortality prediction are reported in **Table S5** with five-fold cross validation. Gradient boosting learning shows the best performance in prediction, recall and AUC evaluation metrics.

The results from the sensitivity analysis excluding NLR and PNI are shown in **Supplementary Appendix 2**. The optimum tree numbers are shown in **Figure S4**. Without NLR and PNI, age became the most important variable for predicting both 30-day and 90-day mortality (**Table S6; Figure S5**) and some evaluation metrics were lower, but others were not affected (**Table S7**). Five-cross validations indicate that the machine learning model maintains comparable prediction performance as the case with NLR and PNI to predict 30-day mortality (precision=0.90, recall=0.89, AUC=0.90) and 90-day mortality (precision=0.91, recall=0.91, AUC=0.90).

## Discussion

The main findings of this study are that 1) PNI, NLR and age in the modified electronic frailty index were the most predictive variables for the short-term mortality outcomes in heart failure patients, and 2) non-linear partial dependence relationships between these predictors and outcomes were observed.

We developed a modified electronic frailty model after incorporating the inflammatory and nutritional indices into the conventional frailty scoring system based on the value of importance of each variable generated from gradient boosting learning model. Compared with multivariate logistic regression and decision tree, gradient boosting learning techniques improved the predictive performance of our frailty model. To enhance mortality prediction by capturing the non-linear pattern within characteristics, we develop an interpretable machine learning model based on gradient boosting machine (31). Machine learning models can be fitted to data individually or combined in an ensemble, resulting in an efficient combination of simple individual learning models that together create a more powerful model (32).

In this study, significant risk factors to predict 30-day and 90-day mortality are efficiently identified with gradient boosting learning model. The obtained rankings of important variables for mortality prediction can be used as an electronic heart failure frailty scoring tool in for clinical use. The efficient identification of partial dependence for predictive variables provides more refined estimation of the likelihood of mortality. For example, effective estimations about patient’s mortality probability based on characteristics of smaller PNI, older age, larger NLR (below 60 or so) and pneumonia. All of these variables were associated with impaired mobility. Of these factors, pneumonia is a common nosocomial condition that also confers a significantly higher risk of 30-day post-admission mortality(33). In addition, we extensively conduct the analysis without PNI and NLR, and the results are provided in **Supplementary Material**. Variable importance ranking for 30-day identifies important variables age, pneumonia, skin ulcer, UTI, Parkinson’s, gout, and male sex to predict 30-day mortality, while variables age, pneumonia, UTI, skin ulcer, Parkinson’s, Carlson score to predict 90-day mortality.

Heart failure has been recognized as predominately a syndrome that affects the geriatric population, with over 50% of incidence and 60% of heart failure-associated mortality occurring in the population over 75 years old (34). Age at diagnosis is also one of the most significant prognostic factors for subsequent survival (35). In our cohort, the median age was 81 years old and the risk of the short-term mortality increased strikingly in those aged over 80. Age was ranked as the most important variable to predict 90-day mortality and the second most important variable for 30-day mortality. In 2011, it was reported that the one-year mortality rates increased sharply from 20% to over 30% in patients 75-84 years, and over 40% in patients aged over 85 years (36). The high prevalence of important risk factors, such as hypertension and ischemic heart disease, leads to the increasing incidence of heart failure in older patients (37). Moreover, the survival outcomes of heart failure are closely related to the unfavorable age-associated changes in cardiovascular structure and function, which compromise cardiac reverse capacity (38). Therefore, it is not surprising to observe the strong prognostic value of age in our frailty model.

The frailty index was based on the concept that frailty is caused by the accumulation of health deficits (39). The frailty state itself is considered as an individual variable that can predict mortality (40), even independently of age in different settings (41). The first electronic frailty index developed by Segal *et al*. was based on the same concept, in which the candidate variables were selected based on their potential correlation with the frailty state rather than mortality directly.^4^ Therefore, the individual variables in the frailty index might not associate well with the mortality outcome, the deficits cumulatively lead to an increased risk of mortality.

The specific value of frailty in heart failure cohort has been examined by many studies. A recent meta-analysis has confirmed the association between the frailty state and the worse clinical outcomes in patients with heart failure (42). Indeed, recent guidelines have recommended the assessment of frailty status in heart failure patients to aid risk stratification (43). The Identification of Senior At Risk (ISAR) scale is another frailty screening tool that can predict 30-day mortality in older patients with acute heart failure (44). Among the current literature, a few studies utilized frailty indices and reached similar conclusions to other studies in which frailty was assessed as a phenotype (45, 46), and there is no consensus which method is more suitable in the cohort of heart failure patients (42). The variables included in the various frailty indices used for heart failure were also largely different. A study in the UK combined the frailty index and nutritional index and found an improved prognostic power compared to the conventional frailty index, suggesting that nutrition and frailty are correlated but also remained as independent prognostic factors (46). No previous study has attempted to incorporate inflammatory measures into frailty indices for heart failure prognosis despite the strong pathophysiological associations between these concepts (47).

### Strength and limitations

To the best of our knowledge, this is the first study incorporating both the inflammatory marker and nutritional index into the conventional frailty index. The indices used in this study, NLR and PNI can be easily calculated and incorporated into the decision-making process in the clinical setting. We utilized a large patient cohort that is homogeneously Chinese from a real-world database and the final frailty score was derived from a machine learning model, which was shown to have better performance than the baseline multivariate logistic regression for mortality prediction. There are some limitations to our study. Firstly, this is a multicenter study conducted in Hong Kong, external validation of our results using data from other databases in other countries are needed. Secondly, our study did not include information on the treatment prescribed during the acute phase and postadmission, which may affect the survival outcomes in patients. Nevertheless, frailty models without adjusting such information are still strong predictors of mortality.(48)

## Conclusions

In this study, we created an electronic frailty index that included comorbidity information, inflammatory and nutritional indices. This was then used for short-term mortality prediction in heart failure. Given that these variables can be determined or calculated automatically, their incorporation into clinical risk scores or prediction rules will facilitate clinicians to perform risk stratification more readily. Further prospective studies are warranted to validate the present model by combining other more comprehensive and complex inflammatory, nutritional and frailty assessment tools to confirm its predictive power for clinical use.

## Data Availability

Data available upon request.

## References

1. Xue QL. The frailty syndrome: definition and natural history. Clin Geriatr Med. 2011 Feb;27(1):1–15.

2. Segal JB, Huang J, Roth DL, Varadhan R. External validation of the claims-based frailty index in the national health and aging trends study cohort. Am J Epidemiol. 2017 Sep 15;186(6):745–747.

3. Fried LP, Tangen CM, Walston J, Newman AB, Hirsch C, Gottdiener J, Seeman T, Tracy R, Kop WJ, Burke G, McBurnie MA, Cardiovascular Health Study Collaborative Research G. Frailty in older adults: evidence for a phenotype. J Gerontol A Biol Sci Med Sci. 2001 Mar;56(3):M146–156.

4. Segal JB, Chang HY, D.Y, Walston JD, Carlson MC, Varadhan R. Development of a Claims-based Frailty Indicator Anchored to a Well-established Frailty Phenotype. Med Care. 2017 Jul;55(7):716–722.

5. Lorenzo-Lopez L, Maseda A, de Labra C, Regueiro-Folgueira L, Rodriguez-Villamil JL, Millan-Calenti JC. Nutritional determinants of frailty in older adults: A systematic review. BMC Geriatr. 2017 May 15;17(1):108.

6. Soysal P, Stubbs B, Lucato P, Luchini C, Solmi M, Peluso R, Sergi G, Isik AT, Manzato E, Maggi S, Maggio M, Prina AM, Cosco TD, Wu YT, Veronese N. Inflammation and frailty in the elderly: A systematic review and meta-analysis. Ageing Res Rev. 2016 Nov;31:1–8.

7. Gastelurrutia P, Lupon J, Altimir S, de Antonio M, Gonzalez B, Cabanes R, Rodriguez M, Urrutia A, Domingo M, Zamora E, Diez C, Coll R, Bayes-Genis A. Fragility is a key determinant of survival in heart failure patients. Int J Cardiol. 2014 Jul 15;175(1):62–66.

8. Denfeld QE, Winters-Stone K, Mudd JO, Gelow JM, Kurdi S, Lee CS. The prevalence of frailty in heart failure: A systematic review and meta-analysis. Int J Cardiol. 2017 Jun 1;236:283–289.

9. McNallan SM, Chamberlain AM, Gerber Y, Singh M, Kane RL, Weston SA, Dunlay SM, Jiang R, Roger VL. Measuring frailty in heart failure: a community perspective. Am Heart J. 2013 Oct;166(4):768–774.

10. Soukoulis V, Dihu JB, Sole M, Anker SD, Cleland J, Fonarow GC, Metra M, Pasini E, Strzelczyk T, Taegtmeyer H, Gheorghiade M. Micronutrient deficiencies an unmet need in heart failure. J Am Coll Cardiol. 2009 Oct 27;54(18):1660–1673.

11. Shirazi LF, Bissett J, Romeo F, Mehta JL. Role of Inflammation in Heart Failure. Curr Atheroscler Rep. 2017 Jun;19(6):27.

12. Westermann D, Lindner D, Kasner M, Zietsch C, Savvatis K, Escher F, von Schlippenbach J, Skurk C, Steendijk P, Riad A, Poller W, Schultheiss HP, Tschope C. Cardiac inflammation contributes to changes in the extracellular matrix in patients with heart failure and normal ejection fraction. Circ Heart Fail. 2011 Jan;4(1):44-52.

13. Paulus WJ, Tschope C. A novel paradigm for heart failure with preserved ejection fraction: comorbidities drive myocardial dysfunction and remodeling through coronary microvascular endothelial inflammation. J Am Coll Cardiol. 2013 Jul 23;62(4):263–271.

14. Van Linthout S, Tschope C. Inflammation - Cause or Consequence of Heart Failure or Both? Curr Heart Fail Rep. 2017 Aug;14(4):251–265.

15. Edelmann F, Holzendorf V, Wachter R, Nolte K, Schmidt AG, Kraigher-Krainer E, Duvinage A, Unkelbach I, Dungen HD, Tschope C, Herrmann-Lingen C, Halle M, Hasenfuss G, Gelbrich G, Stough WG, Pieske BM. Galectin-3 in patients with heart failure with preserved ejection fraction: results from the Aldo-DHF trial. Eur J Heart Fail. 2015 Feb;17(2):214–223.

16. Vasan RS, Sullivan LM, Roubenoff R, Dinarello CA, Harris T, Benjamin EJ, Sawyer DB, Levy D, Wilson PW, D’Agostino RB, Framingham Heart S. Inflammatory markers and risk of heart failure in elderly subjects without prior myocardial infarction: the Framingham Heart Study. Circulation. 2003 Mar 25;107(11):1486–1491.

17. Yildiz A, Yuksel M, Oylumlu M, Polat N, Akil MA, Acet H. The association between the neutrophil/lymphocyte ratio and functional capacity in patients with idiopathic dilated cardiomyopathy. Anatol J Cardiol. 2015 Jan;15(1):13–17.

18. Zapatero A, Barba R, Gonzalez N, Losa JE, Plaza S, Canora J, Marco J. Influence of obesity and malnutrition on acute heart failure. Rev Esp Cardiol (Engl Ed). 2012 May;65(5):421–426.

19. Anker SD, Ponikowski P, Varney S, Chua TP, Clark AL, Webb-Peploe KM, Harrington D, Kox WJ, Poole-Wilson PA, Coats AJ. Wasting as independent risk factor for mortality in chronic heart failure. Lancet. 1997 Apr 12;349(9058):1050–1053.

20. Chien SC, Lo CI, Lin CF, Sung KT, Tsai JP, Huang WH, Yun CH, Hung TC, Lin JL, Liu CY, Hou CJ, Tsai IH, Su CH, Yeh HI, Hung CL. Malnutrition in acute heart failure with preserved ejection fraction: clinical correlates and prognostic implications. ESC Heart Fail. 2019 Oct;6(5):953–964.

21. Cheng YL, Sung SH, Cheng HM, Hsu PF, Guo CY, Yu WC, Chen CH. Prognostic Nutritional Index and the Risk of Mortality in Patients With Acute Heart Failure. J Am Heart Assoc. 2017 Jun 25;6(6).

22. Ueland T, Gullestad L, Nymo SH, Yndestad A, Aukrust P, Askevold ET. Inflammatory cytokines as biomarkers in heart failure. Clin Chim Acta. 2015 Mar 30;443:71–77.

23. Anker SD, Negassa A, Coats AJ, Afzal R, Poole-Wilson PA, Cohn JN, Yusuf S. Prognostic importance of weight loss in chronic heart failure and the effect of treatment with angiotensin-converting-enzyme inhibitors: an observational study. Lancet. 2003 Mar 29;361(9363):1077–1083.

24. Mano A, Fujita K, Uenomachi K, Kazama K, Katabuchi M, Wada K, Terakawa N, Arai K, Hori Y, Hashimoto S, Nakatani T, Kitamura S. Body mass index is a useful predictor of prognosis after left ventricular assist system implantation. J Heart Lung Transplant. 2009 May;28(5):428–433.

25. Desai RJ, Wang SV, Vaduganathan M, Evers T, Schneeweiss S. Comparison of Machine Learning Methods With Traditional Models for Use of Administrative Claims With Electronic Medical Records to Predict Heart Failure Outcomes. JAMA Network Open. 2020;3(1):e1918962–e1918962.

26. Ju C, Lai RWC, Li KHC, Hung JKF, Lai JCL, Ho J, Liu Y, Tsoi MF, Liu T, Cheung BMY, Wong ICK, Tam LS, Tse G. Comparative cardiovascular risk in users versus non-users of xanthine oxidase inhibitors and febuxostat versus allopurinol users. Rheumatology (Oxford). 2019 Dec 24.

27. Lau WC, Chan EW, Cheung CL, Sing CW, Man KK, Lip GY, Siu CW, Lam JK, Lee AC, Wong IC. Association Between Dabigatran vs Warfarin and Risk of Osteoporotic Fractures Among Patients With Nonvalvular Atrial Fibrillation. JAMA. 2017 Mar 21;317(11):1151–1158.

28. Deyo RA, Cherkin DC, Ciol MA. Adapting a clinical comorbidity index for use with ICD-9-CM administrative databases. J Clin Epidemiol. 1992 Jun;45(6):613–619.

29. Llew Mason JB, Peter Bartlett, Marcus Frean Boosting Algorithms as Gradient Descent Advances in neural information processing systems 2000:512–518.

30. Desai RJ, Wang SV, Vaduganathan M, Evers T, Schneeweiss S. Comparison of Machine Learning Methods With Traditional Models for Use of Administrative Claims With Electronic Medical Records to Predict Heart Failure Outcomes. JAMA Netw Open. 2020 Jan 3;3(1):e1918962.

31. Friedman JH. Greedy Function Approximation: A Gradient Boosting Machine. The Annals of Statistics. 2001;29(5):1189–1232.

32. Peter Bühlmann TH. Boosting Algorithms: Regularization, Prediction and Model Fitting. Statistical Science. 2007;22(4):477–505.

33. Kundi H, Wadhera RK, Strom JB, Valsdottir LR, Shen C, Kazi DS, Yeh RW. Association of Frailty With 30-Day Outcomes for Acute Myocardial Infarction, Heart Failure, and Pneumonia Among Elderly Adults. JAMA Cardiol. 2019 Nov 1;4(11):1084–1091.

34. Rich MW. Heart failure in the 21st century: a cardiogeriatric syndrome. J Gerontol A Biol Sci Med Sci. 2001 Feb;56(2):M88–96.

35. Taylor CJ, Ordonez-Mena JM, Roalfe AK, Lay-Flurrie S, Jones NR, Marshall T, Hobbs FDR. Trends in survival after a diagnosis of heart failure in the United Kingdom 2000-2017: population based cohort study. BMJ. 2019 Feb 13;364:223.

36. Chen J, Normand SL, Wang Y, Krumholz HM. National and regional trends in heart failure hospitalization and mortality rates for Medicare beneficiaries, 1998-2008. JAMA. 2011 Oct 19;306(15):1669–1678.

37. Lakatta EG, Levy D. Arterial and cardiac aging: major shareholders in cardiovascular disease enterprises: Part I: aging arteries: a “set up” for vascular disease. Circulation. 2003 Jan 7;107(1):139–146.

38. Lakatta EG, Levy D. Arterial and cardiac aging: major shareholders in cardiovascular disease enterprises: Part II: the aging heart in health: links to heart disease. Circulation. 2003 Jan 21;107(2):346–354.

39. Mitnitski AB, Mogilner AJ, Rockwood K. Accumulation of deficits as a proxy measure of aging. ScientificWorldJournal. 2001 Aug 8;1:323–336.

40. Clegg A, Young J, Iliffe S, Rikkert MO, Rockwood K. Frailty in elderly people. Lancet. 2013 Mar 2;381(9868):752–762.

41. Hewitt J, Carter B, McCarthy K, Pearce L, Law J, Wilson FV, Tay HS, McCormack C, Stechman MJ, Moug SJ, Myint PK. Frailty predicts mortality in all emergency surgical admissions regardless of age. An observational study. Age and Ageing. 2019;48(3):388–394.

42. Zhang Y, Yuan M, Gong M, Tse G, Li G, Liu T. Frailty and Clinical Outcomes in Heart Failure: A Systematic Review and Meta-analysis. J Am Med Dir Assoc. 2018 Nov;19(11):1003–1008 e1001.

43. Díez-Villanueva P, Arizá-Solé A,Vidán MT, Bonanad C, Formiga F, Sanchis J, Martín-Sánchez FJ, Ruiz Ros V, Sanmartín Fernández M, Bueno H, Martínez-Sellés M. Recommendations of the Geriatric Cardiology Section of the Spanish Society of Cardiology for the Assessment of Frailty in Elderly Patients With Heart Disease. Revista Española de Cardiología (English Edition). [10.1016/j.rec.2018.06.035]. 2019;72(1):63–71.

44. Martin-Sanchez FJ, Llopis Garcia G, Gonzalez-Colaco Harmand M, Fernandez Perez C, Gonzalez Del Castillo J, Llorens P, Herrero P, Jacob J, Gil V, Dominguez-Rodriguez A, Rossello X, Miro O, en representacion de los investigadores del Registro OAK, Resto de investigadores del registro OAK. Identification of Senior At Risk scale predicts 30-day mortality among older patients with acute heart failure. Med Intensiva. 2020 Jan - Feb;44(1):9–17.

45. Dunlay SM, Park SJ, Joyce LD, Daly RC, Stulak JM, McNallan SM, Roger VL, Kushwaha SS. Frailty and outcomes after implantation of left ventricular assist device as destination therapy. J Heart Lung Transplant. 2014 Apr;33(4):359–365.

46. Sze S, Zhang J, Pellicori P, Morgan D, Hoye A, Clark AL. Prognostic value of simple frailty and malnutrition screening tools in patients with acute heart failure due to left ventricular systolic dysfunction. Clin Res Cardiol. 2017 Jul;106(7):533-541.

47. Bellumkonda L, Tyrrell D, Hummel SL, Goldstein DR. Pathophysiology of heart failure and frailty: a common inflammatory origin? Aging Cell. 2017 Jun;16(3):444–450.

48. Kojima G, Iliffe S, Walters K. Frailty index as a predictor of mortality: a systematic review and meta-analysis. Age Ageing. 2018 Mar 1;47(2):193–200.

