## Supplementary Appendix 1 for "Derivation of an electronic frailty index for short-term mortality in heart failure: a machine learning approach"

**Table S1.** Variables used in the modified frailty index proposed by Segal et al. (2017), Deyo-Charlson comorbidity index (1992), and our electronic frailty index.

| Segal et al. modified frailty index | Deyo-Charlson comorbidity index | Our electronic frailty index |
| --- | --- | --- |
| 1. Impaired mobility 2. Depression 3. Congestive Heart Failure 4. Parkinson’s disease 5. White race 6. Arthritis (any type) 7. Cognitive impairment 8. Charlson comorbidity index (>0, 0) 9. Stroke 10. Paranoia 11. Chronic skin ulcer 12. Pneumonia 13. Male sex 14. Skin and soft tissue infection 15. Mycoses 16. Age (in 5 year categories) 17. Admission in past 6 months 18. Gout or other crystal-induced arthropathy 19. Falls 20. Musculoskeletal problems 21. Urinary tract infection | 1. Cancer 2. Chronic pulmonary disease 3. Diabetes without complications 4. Congestive heart failure 5. Cerebrovascular disease 6. Dementia 7. Renal disease 8. Peripheral vascular disease 9. Myocardial infarction 10. Diabetes with complications 11. Paraplegia and hemiplegia 12. Connective tissue disease- Rheumatic disease 13. Peptic ulcer disease 14. Mild liver disease 15. Metastatic carcinoma 16. Moderate or severe liver disease 17. HIV/AIDS | 1. Prognostic nutritional index 2. Age 3. Neutrophil-to-lymphocyte ratio 4. Pneumonia 5. Skin ulcer 6. Urinary tract infection 7. Parkinson’s disease 8. Male sex 9. Skin and soft tissue infection 10. Gout 11. Falls 12. Charlson Score>=2 13. Arthritis 14. Mycoses 15. Depression 16. Paranoia |

**Table S2.** Univariate logistic regression for 30-day and 90-day mortality

UTI, urinary tract infection; PNI, prognostic nutritional index; NLR, neutrophil-to-lymphocyte ratio.

|  | **30-day mortality (n=758)** | | **90-day mortality (n=1472)** | |
| --- | --- | --- | --- | --- |
| **Variables** | **Crude Odds Ratio**  **(95% CI)** | **P-Value** | **Crude Odds Ratio**  **(95% CI)** | **P-Value** |
| Male sex | 0.96 (0.83-1.12) | 0.634 | 0.91 (0.82-1.02) | 0.114 |
| Age | 1.04 (1.04-1.05) | **<0.001** | 1.05 (1.04-1.05) | **<0.001** |
| Depression | 0 (0- +$\infty$) | 0.999 | 0.34 (0.04-2.54) | 0.291 |
| Parkinson’s disease | 1.71 (0.88-3.35) | 0.116 | 1.66 (0.97-2.83) | 0.064 |
| Arthritis | 0.87 (0.27-2.83) | 0.816 | 1.07 (0.47-2.42) | 0.872 |
| Paranoid | 0 (0- +$\infty$) | 0.999 | 0 (0- +$\infty$) | 0.999 |
| Skin ulcer | 2.22 (1.33-3.71) | **0.002** | 2.52 (1.68-3.79) | **<0.001** |
| Pneumonia | 2.62 (2.25-3.06) | **<0.001** | 2.36 (2.10-2.67) | **<0.001** |
| Falls | 1.13 (0.69-1.84) | 0.635 | 1.18 (0.82-1.71) | 0.370 |
| Skin and soft tissue infection | 1.07 (0.54-2.14) | 0.839 | 1.05 (0.62-1.77) | 0.868 |
| Mycoses | 1.26 (0.29-5.48) | 0.755 | 0.59 (0.15-2.57) | 0.479 |
| Gouty arthropathy | 1.20 (0.87-1.66) | 0.262 | 1.25 (0.99-1.60) | 0.066 |
| UTI | 1.72 (1.37-2.15) | **<0.001** | 2.03 (1.71-2.40) | **<0.001** |
| Charlson score>=2 | 1.05 (0.90-1.21) | 0.565 | 1.22 (1.09-1.36) | **0.001** |
| NLR | 1.03 (1.02-1.03) | **<0.001** | 1.02 (1.02-1.03) | **<0.001** |
| PNI | 0.94 (0.93-0.95) | **<0.001** | 0.94 (0.93-0.94) | **<0.001** |

**Table S3.** Multivariate logistic regression for 30-day mortality

|  | **Adjust odds ratio**  **(95% CI)** | **Z-value** | **P-value** |
| --- | --- | --- | --- |
| Age | 1.04 (1.03-1.04) | 8.16 | **<0.001** |
| PNI | 0.96 (0.95-0.97) | -6.91 | **<0.001** |
| NLR | 1.01 (1-1.02) | 3.12 | **0.002** |
| Male sex | 1.18 (1-1.39) | 2.00 | **0.046** |
| Depression | 0 (0- +$\infty$) | -0.06 | 0.954 |
| Parkinson’s disease | 1.29 (0.65-2.59) | 0.73 | 0.467 |
| Arthritis | 0.71 (0.21-2.36) | -0.56 | 0.577 |
| Paranoia | 0 (0- +$\infty$) | -0.04 | 0.969 |
| Skin ulcer | 1.76 (1.02-3.04) | 2.04 | **0.041** |
| Pneumonia | 2.23 (1.9-2.62) | 9.68 | **<0.001** |
| Falls | 0.91 (0.55-1.51) | -0.36 | 0.717 |
| Skin and soft tissue infection | 0.77 (0.38-1.58) | -0.71 | 0.476 |
| Mycoses | 0.98 (0.21-4.48) | -0.03 | 0.979 |
| Gout | 0.91 (0.65-1.27) | -0.57 | 0.571 |
| UTI | 1.28 (1.01-1.63) | 2.06 | **0.039** |
| Charlson Score>=2 | 0.98 (0.84-1.15) | -0.26 | 0.792 |

UTI, urinary tract infection; PNI, prognostic nutritional index; NLR, neutrophil-to-lymphocyte ratio.

**Table S4.** Multivariate logistic regression for 90-day mortality

|  | **Adjust odds ratio**  **(95% CI)** | **Z-value** | **P-value** |
| --- | --- | --- | --- |
| Age | 1.05 (1.04-1.05) | 11.73 | **<0.001** |
| PNI | 0.94 (0.93-0.94) | -10.62 | **<0.001** |
| NLR | 1.02 (1.02-1.03) | 1.18 | 0.238 |
| Male sex | 1.14 (1.01-1.29) | 2.09 | **0.036** |
| Depression | 0.33 (0.04-2.58) | -1.06 | 0.288 |
| Parkinson’s disease | 1.19 (0.68-2.09) | 0.62 | 0.532 |
| Arthritis | 0.79 (0.34-1.83) | -0.55 | 0.581 |
| Paranoia | 0 (0- +$\infty$) | -0.07 | 0.948 |
| Skin ulcer | 1.94 (1.25-3.01) | 2.94 | **0.003** |
| Pneumonia | 1.99 (1.75-2.26) | 10.61 | **<0.001** |
| Falls | 0.91 (0.62-1.34) | -0.46 | 0.644 |
| Skin and soft tissue infection | 0.73 (0.42-1.28) | -1.09 | 0.274 |
| Mycoses | 0.46 (0.10-2.10) | -1.00 | 0.316 |
| Gout | 0.93 (0.72-1.20) | -0.55 | 0.582 |
| UTI | 1.5 (1.25-1.80) | 4.38 | **<0.001** |
| Charlson Score>=2 | 1.13 (1.00-1.27) | 1.96 | **0.050** |

UTI, urinary tract infection; PNI, prognostic nutritional index; NLR, neutrophil-to-lymphocyte ratio.

**Table S5.** Five-fold cross validation model performance for 30-day and 90-day mortality prediction

AUC: area under the curve; CI: confidence interval;

| **Model** | **30-day mortality** | | | **90-day mortality** | | |
| --- | --- | --- | --- | --- | --- | --- |
|  | **Precision** | **Recall** | **AUC[95% CI]** | **Precision** | **Recall** | **AUC[95% CI]** |
| Gradient boosting | 0.91 | 0.89 | 0.90 [0.87,0.92] | 0.94 | 0.93 | 0.92 [0.88,0.94] |
| Decision tree | 0.88 | 0.86 | 0.87 [0.85,0.89] | 0.90 | 0.91 | 0.89 [0.86,0.91] |
| Logistic regression | 0.86 | 0.84 | 0.86 [0.83, 0.88] | 0.84 | 0.83 | 0.86 [0.87,0.88] |

AUC, area under the curve.

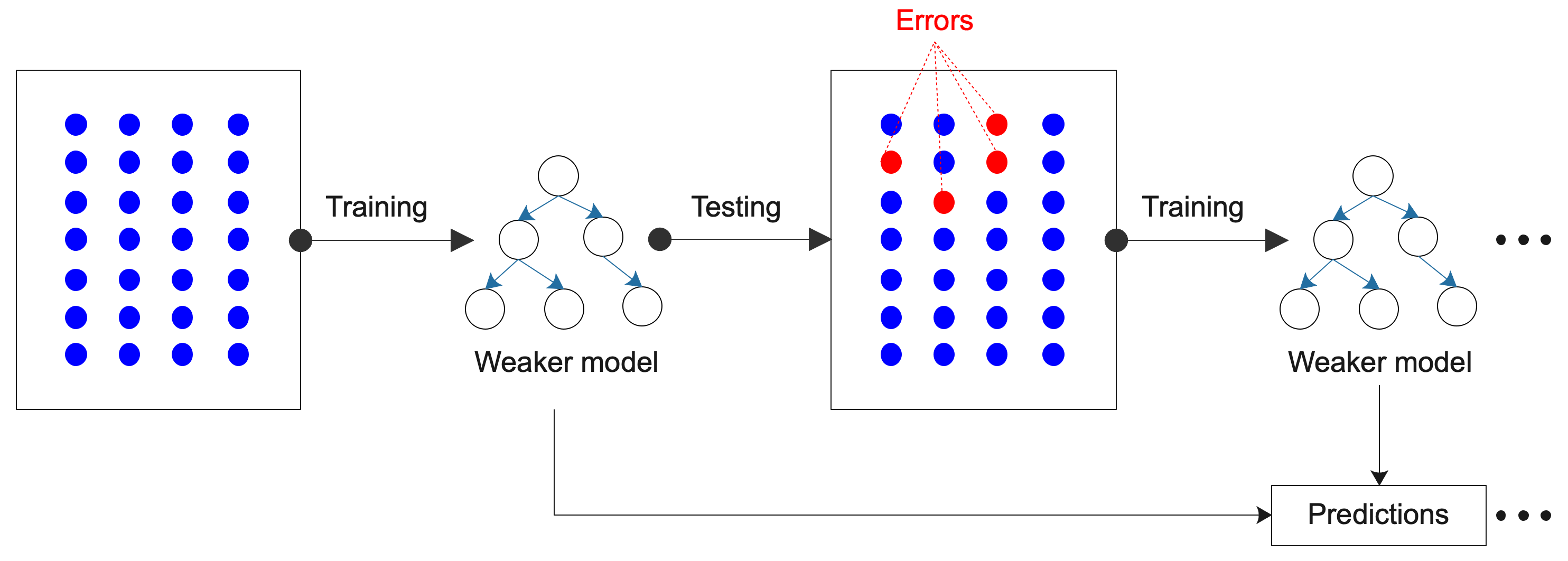

**Figure S1.** Sequential ensemble concept of gradient boosting learning model

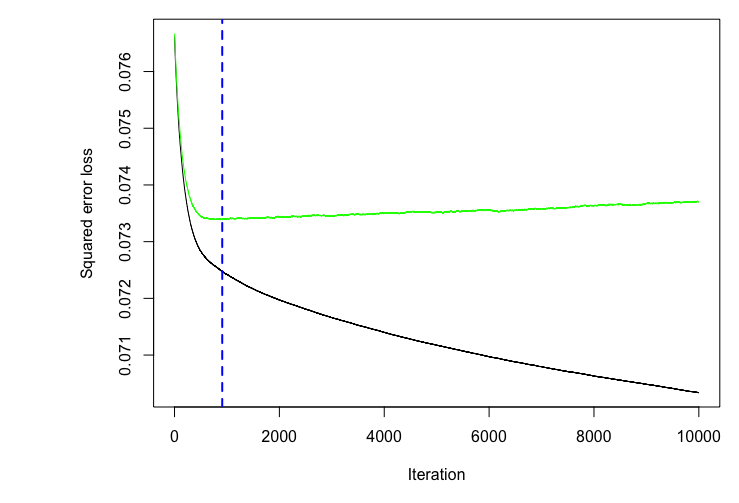

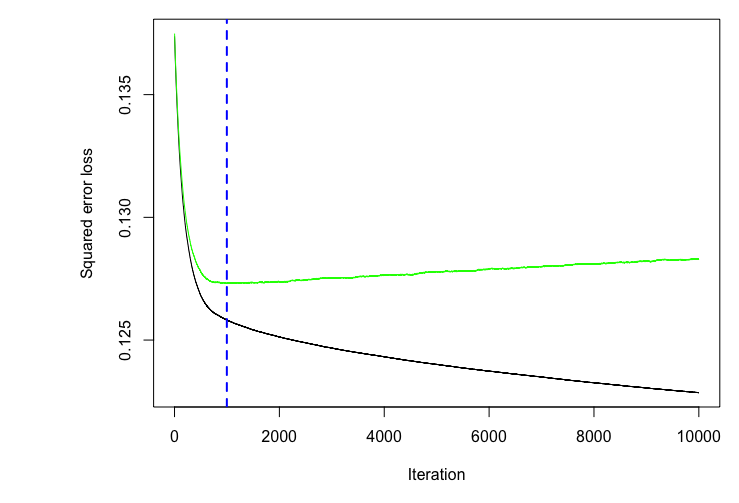

30-day mortality	90-day mortality

**Figure S2.** Optimal iteration tree number of gradient boosting learning model for 30-day and 90-day mortality prediction

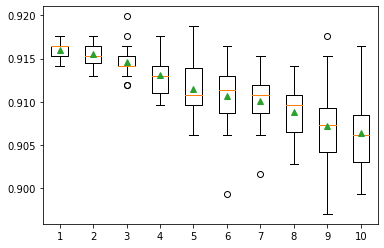

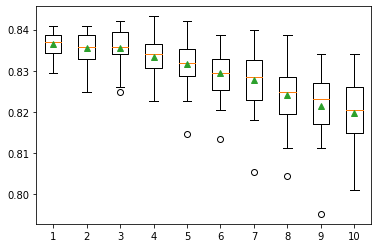

30-day mortality	90-day mortality

The green line shows the practical variation cases with Nelson-Aalen estimator. The iteration stabilizes at around 7.35% when ten thousand trees were employed in the construction of the forest for 30-day mortality prediction, and at around 12.75% for 90-day mortality prediction. The optimal tree number was set according to the dashed blue line. The black line shows the ideal expected OOB error rate variation with tree number variation.

**Figure S3.** Tree in-depth parameter selection for 30-day and 90-day mortality prediction
