## Supplementary Appendix 2 for "Derivation of an electronic frailty index for short-term mortality in heart failure: a machine learning approach"

**Supplementary Appendix 2: Sensitivity Analysis by excluding NLR and PNI**

**Table S6.** Variable importance for 30-day and 90-day mortality prediction with gradient boosting learning without NLR and PNI

| **30-day mortality** | | **90-day mortality** | |
| --- | --- | --- | --- |
| **Variable** | **Importance** | **Variable** | **Importance** |
| Age | 74.26 | Age | 70.79 |
| Pneumonia | 10.31 | Pneumonia | 11.25 |
| Skin.ulcer | 4.37 | UTI | 4.33 |
| UTI | 2.71 | Skin.ulcer | 3.67 |
| Parkinson’s | 2.56 | Parkinson’s | 1.67 |
| Male.sex | 1.36 | Charlson.Score>=2 | 1.62 |
| Gout | 1.22 | Male.sex | 1.61 |
| Falls | 1.03 | Falls | 1.44 |
| Soft.infection | 0.82 | Arthritis | 1.35 |
| Charlson.Score>=2 | 0.80 | Gout | 1.32 |
| Arthritis | 0.52 | Soft.infection | 0.76 |
| Mycoses | 0.05 | Mycoses | 0.14 |
| Depression | 0 | Depression | 0.07 |
| Paranoia | 0 | Paranoia | 0 |

UTI, urinary tract infection.

**Table S7.** Five-fold cross validation model performance for 30-day and 90-day mortality prediction without NLR and PNI

AUC: area under the curve; CI: confidence interval;

| **Model** | **30-day mortality** | | | **90-day mortality** | | |
| --- | --- | --- | --- | --- | --- | --- |
|  | **Precision** | **Recall** | **AUC [95% CI]** | **Precision** | **Recall** | **AUC [95% CI]** |
| Gradient boosting | 0.90 | 0.89 | 0.90[0.87,0.92] | 0.91 | 0.91 | 0.90[0.88,0.93] |
| Decision tree | 0.89 | 0.86 | 0.87[0.85,0.89] | 0.89 | 0.91 | 0.88[0.86,0.92] |
| Logistic regression | 0.86 | 0.84 | 0.86[0.81,0.88] | 0.82 | 0.82 | 0.86[0.82,0.87] |

AUC, area under the curve.

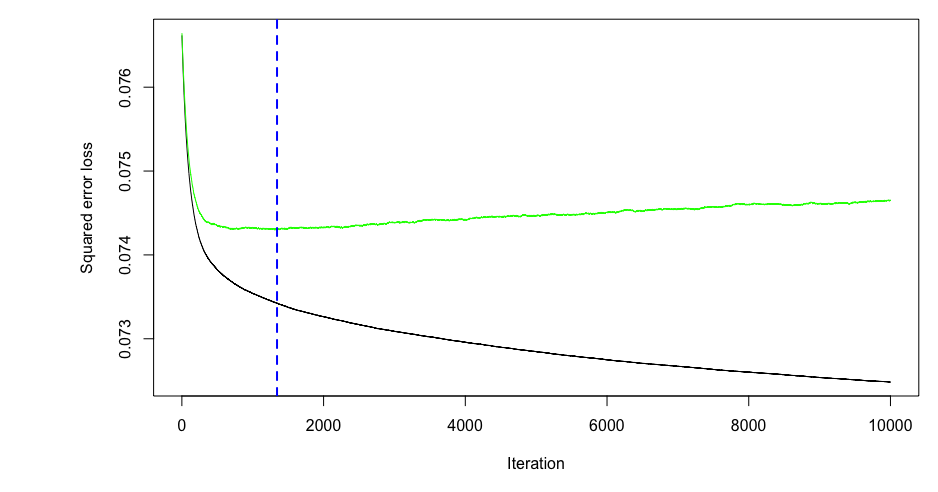

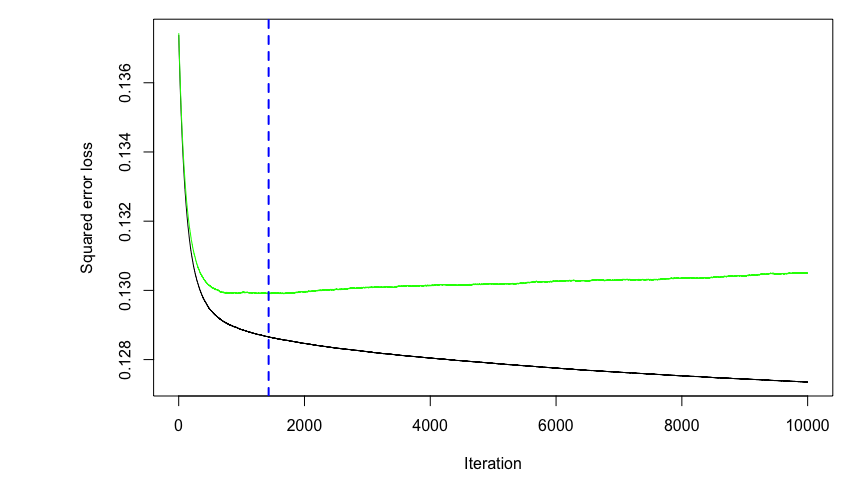

30-day mortality	90-day mortality

**Figure S4**. Optimal iteration tree number of gradient boosting learning model for 30-day and 90-day mortality prediction without NLR and PNI

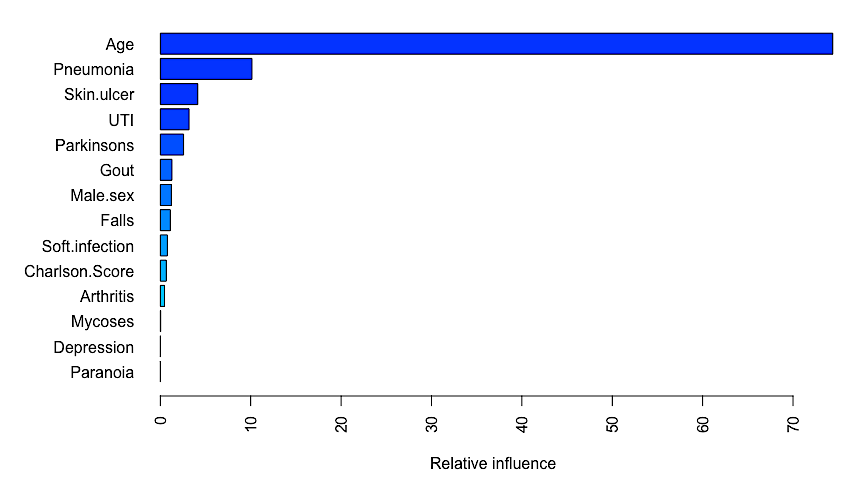

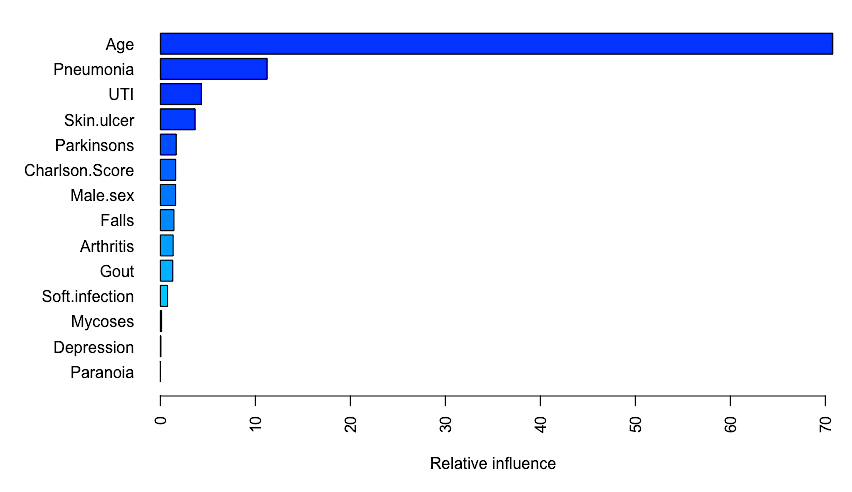

**Figure S5**. Variable importance ranking for 30-day and 90-day mortality prediction without NLR and PNI
